# Comparison of PU.1 genomic binding across immune cells reveals cell type-specific roles in autoimmune disease

**DOI:** 10.64898/2026.01.26.26344838

**Authors:** David A. Lewis, Molly S. Shook, Lucinda P. Lawson, Sreeja Parameswaran, Xiaoting Chen, Lee E. Edsall, Omer A. Donmez, Cailing Yin, Diego A. Rosado-Tristani, Andrew VonHandorf, Phillip J. Dexheimer, Carmy R. Forney, Jennifer M. Felton, Arame A. Diouf, Katelyn A. Dunn, David F. Smith, Marc E. Rothenberg, Jason S. Seidman, Ty D. Troutman, Christopher K. Glass, Stephen N. Waggoner, Matthew T. Weirauch, Leah C. Kottyan

## Abstract

Transcription factors play key roles in cellular biology. Their genomic binding events are enriched at disease- and trait-associated genetic risk loci in particular cellular contexts. To examine this phenomenon in depth, we constructed a PU.1 (SPI1) binding atlas by uniformly processing 260 PU.1 ChIP-seq datasets spanning many immune cell types. Comparison of ChIP-seq peaks across eight major immune cell types identifies shared and cell type-specific PU.1 binding events. DNA sequence analyses reveals context-specific binding shaped by both canonical PU.1 motifs and motifs from partner transcription factor families. Integration of this atlas with genome-wide association studies of blood cell traits and immune-mediated diseases reveals strong enrichment for PU.1 binding events at genetic risk loci and extensive genotype-dependent PU.1 genomic occupancy. We identify cellular contexts in which PU.1 enrichment is most pronounced, including at autoimmune disease loci within EBV-positive B cells. Together, these results define cellular, infectious, and genetic contexts of PU.1 binding that help connect noncoding variation to human phenotypes.

## INTRODUCTION

Genetic association studies have identified thousands of loci that influence blood cell traits and immune-mediated diseases, with most genetic risk variants mapping to noncoding regulatory regions. Interpreting these associations requires understanding how regulatory variants act in specific cellular, environmental, and molecular contexts. Often these mechanisms involve the alteration of transcription factor (TF) occupancy at cis-regulatory elements. TF-centered analyses have shown that the binding of specific TFs can be concentrated across many loci for a given phenotype, nominating shared regulatory mechanisms that operate across the genome. For example, GATA3 occupies over one-third of breast cancer risk loci in a breast cancer-derived cell line, illustrating how TF maps can reveal potential disease mechanisms^1^.

PU.1, encoded by *SPI1*, is an Ets-family transcription factor with central and pleiotropic roles in hematopoiesis. PU.1 is required for myeloid and dendritic-cell development and contributes to B cell fate specification. Likewise, PU.1 shapes developmental potential and delays lineage commitment in early T cell precursors^2^. The activity of PU.1 is highly dose-sensitive, with graded PU.1 expression influencing lineage outcomes through antagonistic interactions with factors such as GATA1^3^. At the *SPI1* locus itself, recent studies have mapped cis-regulatory elements and chromatin architecture that control PU.1 expression across human blood lineages. These efforts revealed a myeloid-biased super-enhancer cluster and an insulated chromatin neighborhood that support lineage-specific enhancer–promoter interactions^4^. Collectively, this work establishes PU.1 as a lineage-defining, context-dependent TF whose own regulation and subsequent downstream binding events are tightly linked to hematopoietic cell identity.

In addition to recognizing a short DNA binding site (core: 5′-GGAA-3′ with additional flanking bases), PU.1 can act either as a pioneer factor that opens chromatin or as a collaboration-dependent TF binding partner. In the latter role, PU.1 forms complexes with activation-induced factors like NF-κB and lineage-restricted cofactors such as C/EBP and IRF family members^2^. Composite DNA binding sites and protein–protein interactions enable PU.1 to integrate local sequence context with cofactor availability, thereby producing distinct binding landscapes in different cell types^5^. For example, during early B cell development, PU.1 dimerizes with IRF4 or IRF8 and binds Ets-IRF composite elements (EICE) to regulate the rearrangement of immunoglobulin light chain genes^6^. In myeloid cells, direct interactions between PU.1 and GATA proteins inhibit myeloid-specific gene activation by blocking PU.1 binding to c-Jun, an important co-activator. Lineage-specifying interactions between PU.1 and several other hematopoietic regulators, including C/EBPa^7, 8^ and RUNX1^9-11^, have been well documented. Classic biochemical studies show that the transcriptional activity of PU.1 can be modulated by direct interactions with lineage-specifying factors such as GATA1, independent of the expression level of PU.1, enabling cross-antagonism between competing differentiation programs^3, 12^. Despite these insights, there exists no systematic view of how PU.1 binding is partitioned into shared and cell type-specific components across human blood lineages. Furthermore, the relationship between context-specific PU.1 genomic occupancy and cofactor binding motifs, composite motifs, and human genetic variation remains incompletely defined.

Recent TF-centered genomic studies have begun to link variation in PU.1 occupancy to blood traits and immune disease. Our own work demonstrated that PU.1 in B cells is enriched at autoimmune and inflammatory bowel disease genetic risk loci^1, 13^. In primary human neutrophils, PU.1 binding across nearly one hundred individuals demonstrated extensive genotype-dependent binding. This work showed that genetic variants affecting PU.1 binding alter chromatin state, enhancer–promoter contacts, and gene expression at loci associated with blood cell counts and autoimmune diseases^14^. In another study, Jeong and Bulyk used lymphoblastoid cell lines to analyze variants with allele-dependent PU.1 binding. Colocalization analysis with GWAS studies of blood cell traits pinpointed dozens of genetic loci where variation in PU.1 motif strength and occupancy likely mediates genetic effects on blood traits^15^. These studies demonstrate that PU.1 occupancy can serve as a powerful lens for prioritizing causal variants at individual GWAS loci and for tracing variant-to-function mechanisms in specific cell types. However, they focus on limited cellular contexts, leaving unresolved how PU.1 binding variation is organized across hematopoietic lineages, and how infection-related or cell state–specific contexts modulate the contribution of PU.1 to genetic risk.

Infection-related and cell state–specific contexts are forms of environmental context that can substantially change how TFs interact with human DNA within the cell. For example, B cells infected with Epstein–Barr virus (EBV) exhibit distinct transcriptional programs compared to uninfected B cells, reshaping both viral and host TF binding at autoimmune risk loci. Our previous work has shown that the EBV transactivating protein EBNA2 anchors gene–environment interactions by occupying many autoimmune disease loci and co-localizing with human TFs such as PU.1, showing that viral infection can rewire TF occupancy patterns at genetically susceptible regions compared to cells without EBV transformation^1, 16, 17^. Given the central role of PU.1 in B cell and myeloid biology, and the established role of EBV in multiple immune-mediated diseases^18^, understanding how the environmental context of EBV status modifies PU.1 binding at disease-associated loci is vital.

Here, we take a TF-centered, cross-context approach to systematically study PU.1 binding in human immune cells. We compile and uniformly process 260 human PU.1 ChIP-seq datasets spanning diverse blood cell types and states. We construct a consensus PU.1 binding atlas encompassing shared and cell type-specific genomic interactions. Using this atlas, we characterize the DNA sequence architecture of these interactions through TF binding motif analyses. We then integrate this atlas with blood cell count and immune-mediated disease GWAS datasets to estimate the enrichment of PU.1 binding at genetic risk loci in different cellular contexts. Finally, leveraging datasets with available genotype information, we discover extensive genotype-dependent PU.1 occupancy at loci associated with these blood cell traits and immune diseases. By jointly analyzing cellular, infection, and genetic contexts, our study provides a comprehensive view of context-specific PU.1 binding in human blood cells, illuminating mechanisms linking noncoding variants to hematologic and immune phenotypes.

## RESULTS

### Construction of a comprehensive PU.1 binding atlas across human hematopoietic lineages

To generate a comprehensive atlas of human PU.1 occupancy, we assembled 296 ChIP–seq datasets, including 253 experiments from publicly available resources and 43 new PU.1 ChIP-seq experiments designed to assess cell types for which no data were available. These data span major hematopoietic populations and cell lines, including large-scale efforts profiling PU.1 in B cells and neutrophils^14, 15^. Each dataset was categorized as one of ten immune cell types: stem cells, myeloid progenitor cells, neutrophils, monocytes, eosinophils, dendritic cells, macrophages, T cells, B cells, or NK cells **(Supplementary Data 1, Fig. 1a)**. Data were uniformly reprocessed using computational pipelines implementing the ENCODE consortium ChIP-seq standards (see Methods) ^19-21^. After stringent quality control incorporating the number of non-duplicate mapped reads, peak counts, and enrichment of the PU.1 motif, 36 datasets were excluded. The remaining high-confidence set of 260 experiments was used for downstream analyses. A substantial fraction of the data was derived from B cell lines, which were further stratified into EBV-positive and EBV-negative lines to capture the effects of virus infection. Notably, despite multiple experimental attempts using protocols and reagents that yielded high-quality data in other contexts, we were unable to generate PU.1 ChIP-seq datasets from T cells or NK cells that passed quality thresholds, and therefore these lineages are not represented in the atlas. Indeed, we did not observe detectable expression of PU.1 in a variety of primary human NK cells or NK cell lines, including after stimulation with pro-inflammatory cytokines (data not shown).

**Figure 1.**
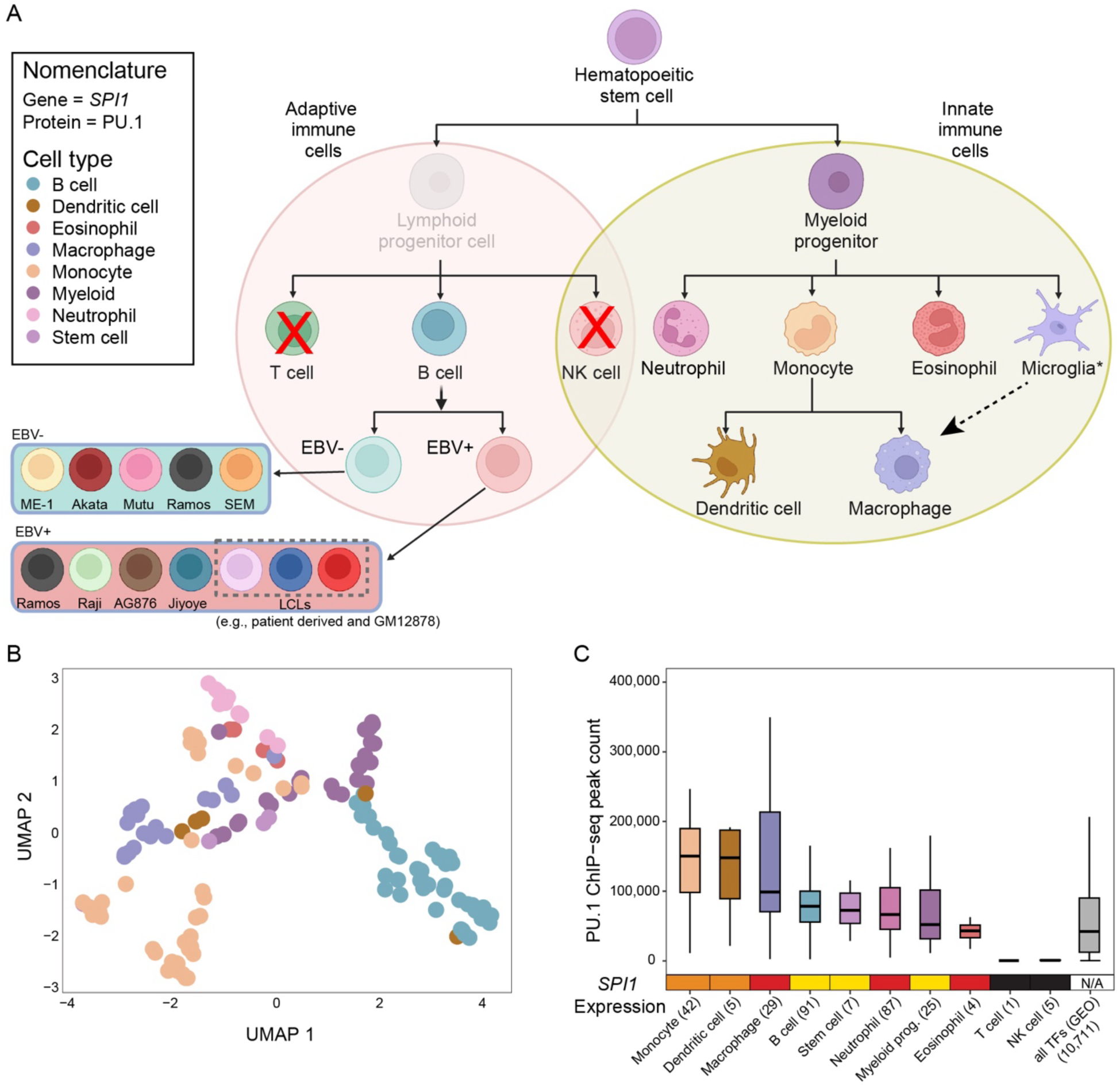
Overview of PU.1 ChIP-seq datasets. A. Cells with available PU.1 ChIP-seq data are presented in the context of hematopoiesis. Red X’s indicate cell types without high quality PU.1 ChIP-seq data despite application of experimental methods that resulted in robust results in other cell types. No data are available for lymphoid progenitor cells. Dashed arrow indicates the fact that microglia physiologically develop from macrophages but were derived from myeloid progenitors for the datasets in this study. PU.1 datasets were obtained from B cells that were either Epstein-Barr virus (EBV) positive or negative (lower left). B. Relationship of PU.1 ChIP-seq peaks across cell types. Each point represents a PU.1 ChIP-seq experiment. Experiments are clustered using UMAP based on read counts within each peak. Datasets are colored based on cell type. C. Top: Distribution of PU.1 ChIP-seq peak counts across cell types. Each data point represents a single ChIP-seq experiment, with the distribution summarized as a box plot. The grey box on the right indicates peak counts across public ChIP-seq datasets for 296 unique DNA binding transcription factors (TFs). The number of datasets for each cell type is indicated in parentheses. The top, middle, and bottom horizontal lines within each box represent the upper quartile, median, and lower quartile, respectively. The whiskers extend from the boxes to the largest or smallest value no further than 1.5 times the interquartile range from the boxes. Values outside of the box and whiskers range are plotted as individual points. Bottom: The expression level of *SPI1* (PU.1 gene) was categorized as high (red), intermediate (orange), low (yellow), or not expressed (black) based on public datasets (see Methods).

To assess how PU.1 binding patterns relate to cellular identity and potential technical variables, we performed dimensionality reduction analysis of peak read counts using Uniform Manifold Approximation and Projection (UMAP). PU.1 ChIP-seq experiments clustered primarily by immune cell type **(Fig. 1b)**, rather than by antibody, study of origin, or total peak number **(Supplementary Fig. 1)**. For example, B cells are entirely located in the upper left quadrant of the UMAP despite the datasets being associated with different antibodies, studies, and peak counts. This observation indicates that biological context is the dominant driver of variation in PU.1 occupancy across the atlas.

We next compiled PU.1 gene expression data from multiple immune expression atlases, including the Human Protein Atlas^22^, the Immunological Genome Project^23^, and the Database of Immune Cell eQTLs^24^. Given differences in normalization across platforms, we qualitatively categorized *SPI1* expression as high, intermediate, or low in each cell type **(Fig. 1c**, *SPI1* expression). These qualitative expression levels were broadly concordant across data sources and aligned with the absence of measurable PU.1 binding in T cells and NK cells, providing orthogonal support that the atlas captures expected lineage-specific differences in expression and PU.1 binding. Importantly, we did not observe linear correlation between PU.1 gene expression levels and PU.1 ChIP-seq peak counts **(Fig. 1c)**, suggesting that factors beyond PU.1 expression level (such as chromatin accessibility and cofactor availability) impact the magnitude of genomic PU.1 binding within specific immune cell types.

### Shared and cell type–specific characteristics of the PU.1 binding landscape

To quantify the extent to which PU.1 binding is shared versus cell type-specific, we compared PU.1 ChIP-seq peaks across immune cell types and classified each peak as cell type–specific (present in a single cell type) or present in two or more cell types (multiple) **(Fig. 2a)**. The absolute number of cell type-specific peaks varied substantially by lineage and was largely driven by the number of PU.1 ChIP-seq datasets available for each cell type **(Fig. 2a)**. We next examined the proportion of specific and broadly shared peaks within each lineage **(Fig. 2b)**. Across cell types, we identified a core set of PU.1 peaks that were shared across seven or more cell types and accounted for at least ∼20% of the peaks in each individual cell type (henceforth referred to as the “shared” peak set). This observation is consistent with a common PU.1 regulatory sub-network that is broadly deployed across hematopoietic lineages.

**Figure 2.**
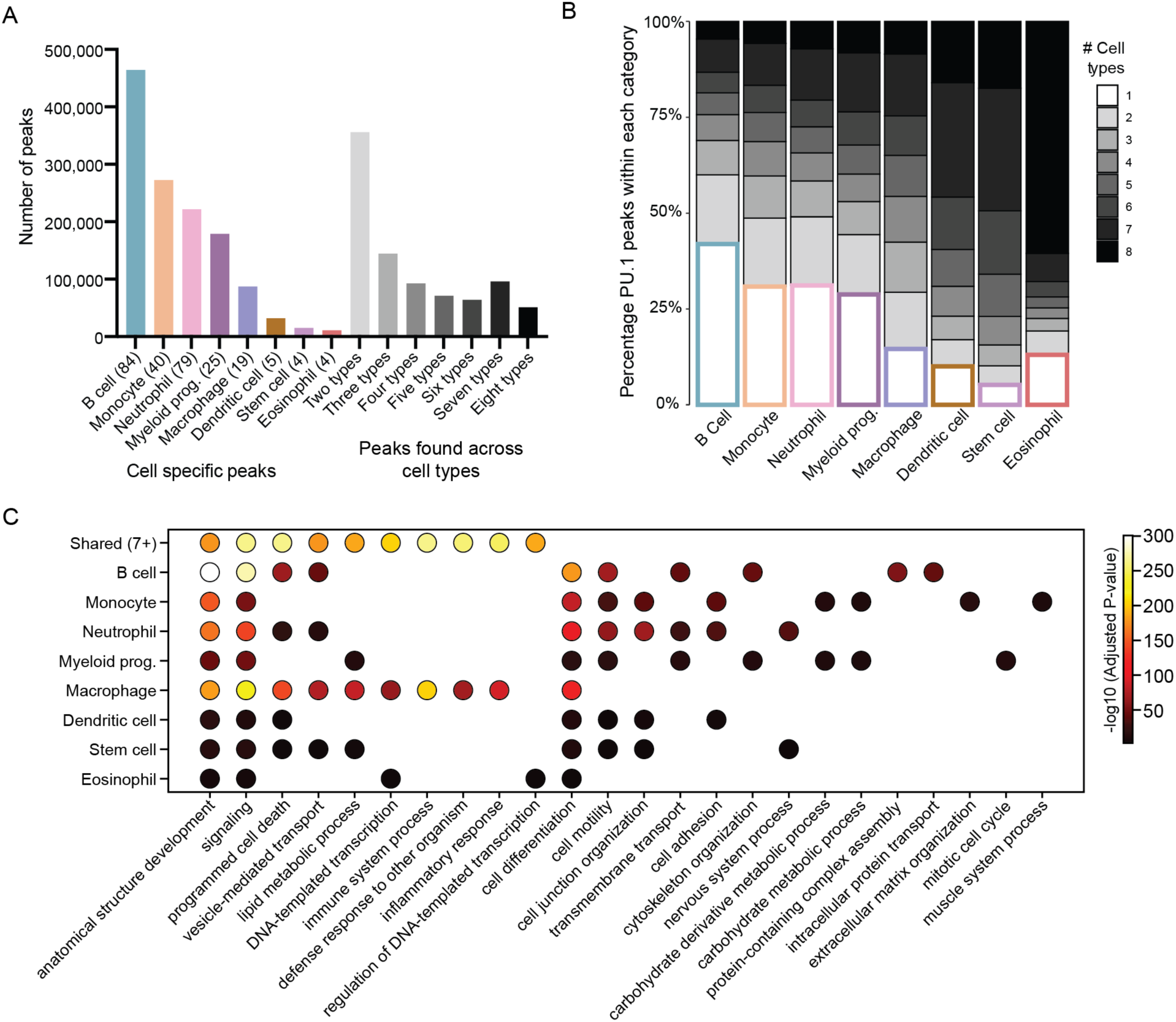
PU.1 ChIP-seq peak comparisons across cell types. Individual PU.1 ChIP-seq datasets were combined within each cell type. Peaks within cell types were further categorized as cell specific or shared. A. Number of non-overlapping PU.1 ChIP-seq peaks within each category. The number of datasets for each cell type is indicated within parentheses after the name. B. Percentage of PU.1 peaks within each category. C. Top ten gene ontology terms enriched for each category of PU.1 peaks. Peaks were mapped to the two nearest genes and pathways enrichment for GO Biological Processes were identified using GREAT. Pathways were restricted to GO Slim terms.

To investigate the biological processes associated with shared versus cell type-specific PU.1 occupancy, we used the GREAT method^25^ to link PU.1 peaks to nearby genes and perform pathway enrichment analyses **(Supplementary Data 2, Fig. 2c)**. This analysis revealed that some pathways are unique to the shared peak set, while others are specific to certain cells. For example, the shared PU.1 peak set was not enriched for cell differentiation, whereas each cell type-specific peak set showed strong enrichment for this pathway. This suggests that lineage-restricted PU.1 binding is preferentially found at regulatory elements involved in cell type-specific differentiation. Additional enrichments in cell type-specific sets included processes such as motility, adhesion, and cytoskeleton organization, highlighting distinct effector programs encoded within PU.1 landscapes of individual immune lineages. In contrast, shared PU.1 peaks were enriched for broadly acting processes, including cell death, transcription, and immune system functions. While some cell type-specific sets also showed these broad enrichments, they included additional cell-specific functions absent from this shared set.

### DNA sequence and composite elements underlying PU.1 binding across immune lineages

We next asked how DNA sequence features and partner transcription-factor motifs shape PU.1 binding across the atlas. As expected, the canonical PU.1 motif was the top enriched motif in every individual dataset that passed our quality control assessment **(Supplementary Data 1)**. When we examined motif enrichment within the shared PU.1 peak set, we found that >95% of the top 100 enriched motifs belonged to the Ets family, which includes PU.1, indicating that PU.1 occupancy at PU.1 sites shared across cell types is predominantly organized around PU.1 binding sites (**Supplementary Data 3, Fig. 3a**). Within the cell type-specific peak sets, however, the relative contribution of Ets motifs varied by lineage. Macrophage-, stem cell-, and B cell-specific peak sets showed a substantially higher proportion of Ets motifs among their top enriched motifs than other cell types. Monocyte- and neutrophil-specific PU.1 peaks showed robust additional enrichment for bZIP motifs. Consistent with prior work in myeloid progenitors, progenitor-specific PU.1 peaks were strongly enriched for GATA motifs^3, 26^. Dendritic cell- and eosinophil-specific peak sets showed a distinctive motif signature, with a prominent contribution from AP-2 and C2H2 zinc finger motifs, highlighting cell type–specific cofactors within the broader PU.1 binding landscape.

**Figure 3.**
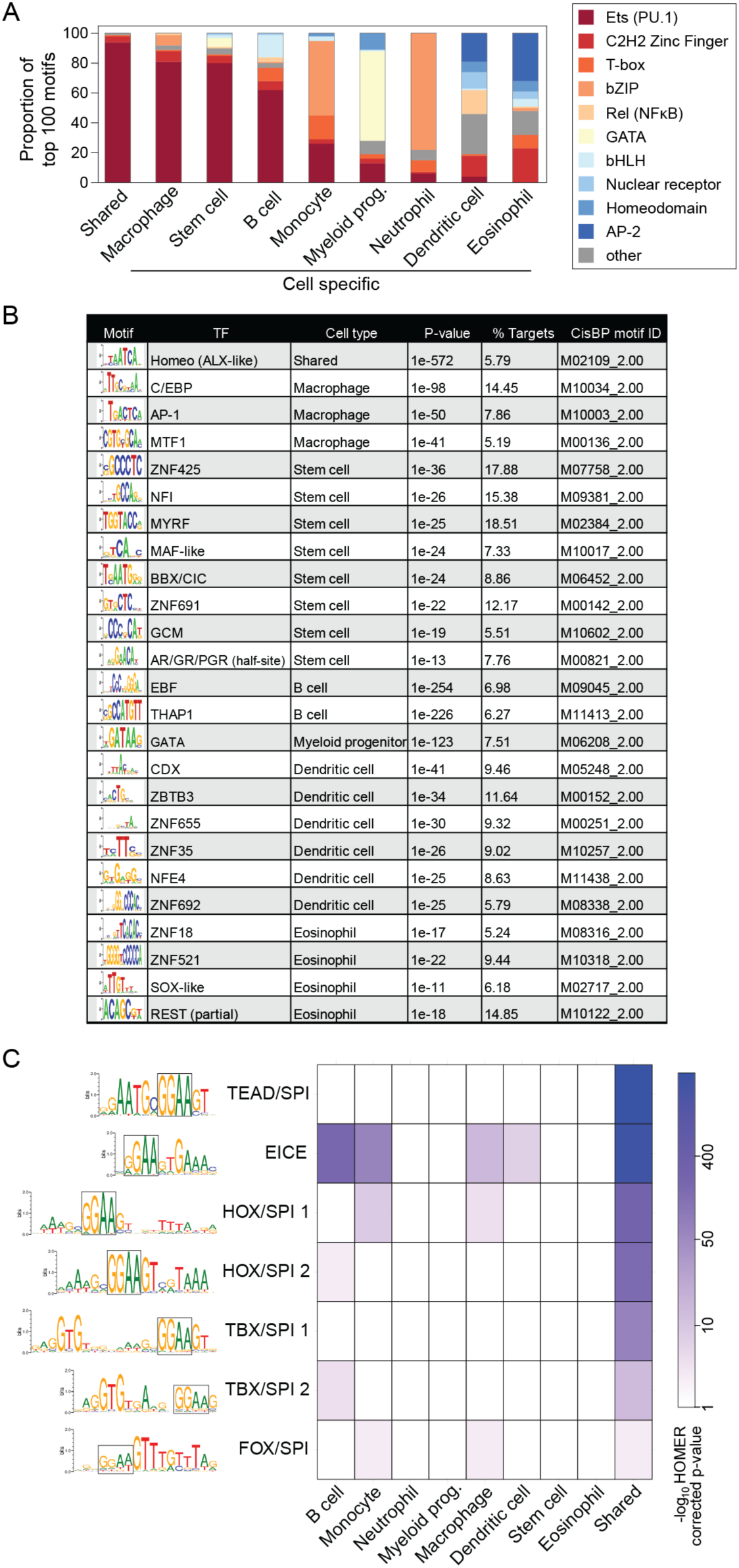
Transcription factor binding site motif enrichment within PU.1 peak categories. A. Motif enrichment within each peak category. Motifs are categorized by TF family (colors). The proportions of the top 100 most significant motifs within each TF family are indicated. B. *De novo* motif enrichment results within each peak category. Non-PU.1 motifs are shown for each peak category that pass the following thresholds: significant HOMER *de novo* p-value (P<0.01), at least 1.2-fold enrichment relative to background, and present in at least 5% of PU.1 peaks. C. Composite motif enrichment within each peak category. A set of nine composite motifs involving PU.1 were compiled (see Methods). The core PU.1 motif is boxed within the sequence logos. Motif enrichment was performed within each peak category using HOMER. Values indicate -log_10_ corrected p-values.

We next sought to more deeply characterize the set of TFs that might collaborate with PU.1 in a cell-specific manner. To this end, we performed HOMER de novo TF motif enrichment analysis on the cell-specific peaks^27^. Figure 3b depicts the most strongly enriched non-PU.1 de novo motifs obtained for each cell-specific peak set. Of note, this analysis identified many of the same TFs found by the known motif analysis (e.g., GATA in myeloid progenitors). It also identified additional possible PU.1 partners, including known relationships such as C/EBP and AP-1 in macrophages^28^ and EBF in B cells^27^, along with new predictions such as THAP1 in B cells.

We next examined composite TF binding motifs containing experimentally validated binding sites for PU.1 together with a cooperating TF. To this end, we queried nine available composite motifs, including those representing the well-characterized PU.1/IRF element (EICE)^27^, along with relatively uncharacterized PU.1 composite binding motifs containing TBX, TEAD, HOX, and FOX family members^29^ (**Supplementary Data 4, Fig. 3b**). Shared PU.1 peaks were enriched for all composite motif classes, with the strongest enrichment observed for TEAD/SPI and EICE motifs. The prominence of TEAD/SPI and EICE elements suggests that PU.1 leverages these partnerships to establish a widely deployed regulatory scaffold that might operate across immune lineages. As expected, B cell–specific PU.1 peaks showed strong enrichment for EICE elements, consistent with established PU.1–IRF cooperativity in B-lineage cells^6^, together with HOX/SPI and TBX/SPI motifs. Monocyte- and macrophage-specific PU.1 peaks were enriched for EICE, HOX/SPI, and FOX/SPI motifs, whereas dendritic cell–specific peaks showed selective enrichment for EICE motifs. Together, these analyses reveal that while Ets motifs are a universal feature of PU.1 binding, cell type–specific PU.1 occupancy is further shaped by distinct repertoires of partner motifs, with IRF/SPI and TEAD/SPI interactions emerging as particularly influential components of PU.1-mediated regulation.

### Cell type–specific enrichment of PU.1 binding at blood cell trait and immune disease risk loci

We and others have previously observed that PU.1 binding is enriched at loci associated with blood cell traits and immune-mediated diseases^1, 15^. To systematically assess the cell-type specificity of these enrichments, we used the RELI algorithm to quantify the overlap between PU.1 ChIP-seq datasets and genome-wide association study (GWAS) loci for blood cell traits and disease phenotypes curated in the GWAS Catalog^1, 30^ (**Supplementary Data 5** and see Methods). Using a meta-analysis framework that aggregated signal within each lineage, we first examined PU.1 enrichment at loci associated with white blood cell traits (**Fig. 4a**, left). PU.1 binding in eosinophils was most strongly enriched at loci associated with eosinophil counts, while monocyte and macrophage PU.1 datasets were strongly enriched at loci associated with monocyte counts. Similarly, neutrophil PU.1 datasets showed strong enrichment at loci associated with neutrophil and granulocyte counts. Thus, the cell-type origin of the PU.1 ChIP–seq experiment closely aligned with the lineage specificity of blood cell trait associations.

**Figure 4.**
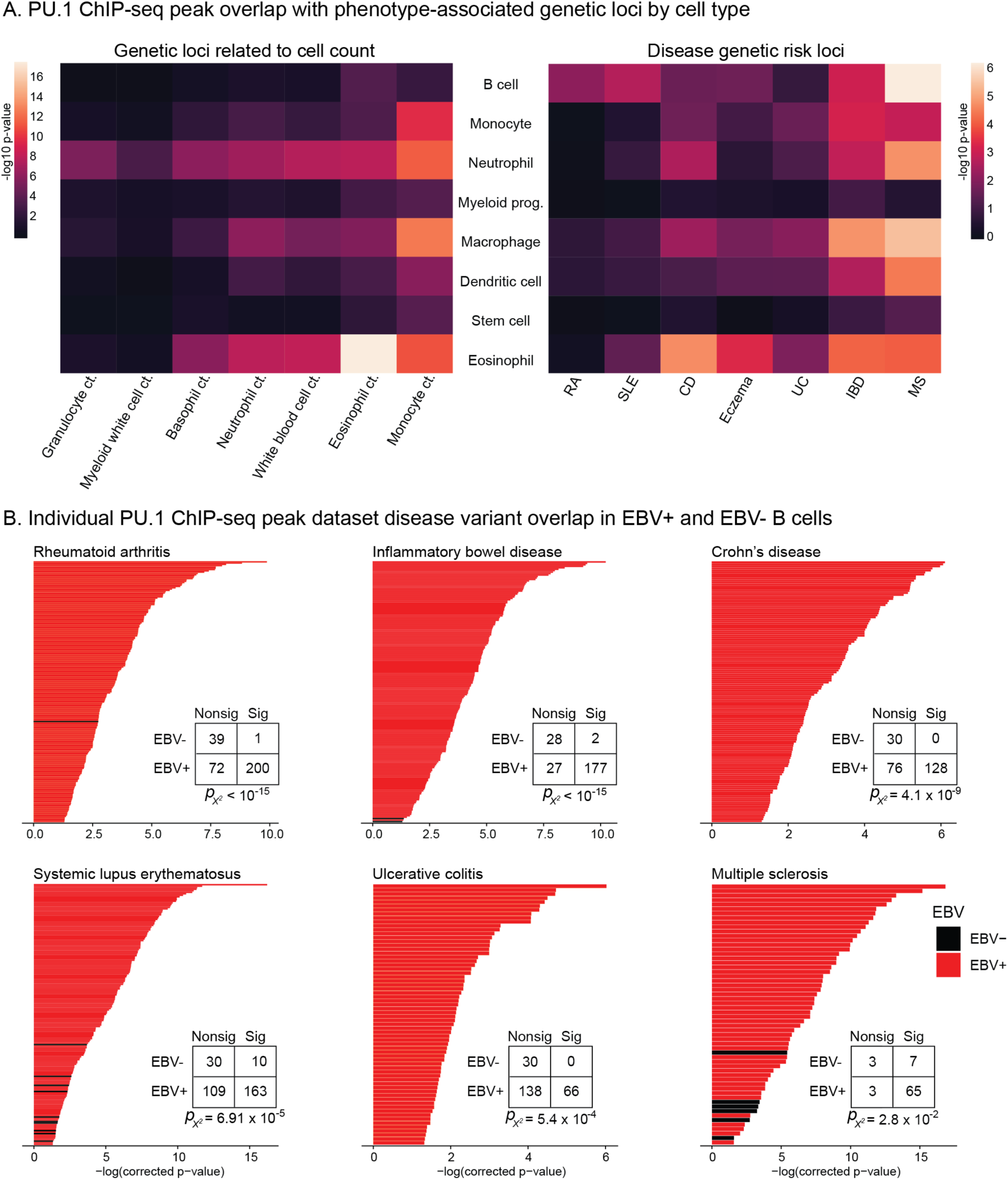
Intersection of PU.1 binding events with phenotype-associated genetic loci. A. PU.1 ChIP-seq peak overlap with phenotype-associated genetic loci by cell type. All phenotypes with at least one meta-p-value result less than 10^-3^ are included. Meta-p-values were calculated by combining individual RELI p-values across datasets obtained from the same cell type (see Methods). Left: Meta-p-values indicating PU.1 ChIP-seq peak overlap with cell count genetic loci, by cell type. Right: Meta-p-values for disease genetic risk loci, by cell type. B. Individual PU.1 ChIP-seq dataset disease variant overlap in EBV+ and EBV- B cells. RELI p-values for EBV+ (red bars) and EBV- (black bars) B cells for individual PU.1 ChIP-seq datasets. Only datasets with RELI p-values less than 0.05 are graphed. Contingency tables show the number of EBV+ and EBV- datasets with RELI p-values greater than 0.05 (“Nonsig”) and less than 0.05 (“Sig”). The Chi Squared p-values represent the enrichment for EBV+ B cells. The total number of datasets differs across diseases based on the number of ancestries with available genetic association data. Disease abbreviations: RA, rheumatoid arthritis; SLE, systemic lupus erythematosus; CD, Crohn’s disease; UC, ulcerative colitis; IBD, inflammatory bowel disease; MS, multiple sclerosis.

Applying the same meta-analytic approach to immune-mediated disease GWAS, we identified seven disease phenotypes with significant PU.1 enrichment in at least one cell type (**Fig. 4a, right**). PU.1 binding in macrophages and eosinophils was most strongly enriched at inflammatory bowel disease (IBD) risk loci, with PU.1 occupancy in eosinophils particularly enriched at Crohn’s disease loci. Eosinophil PU.1 binding was also enriched at loci associated with eczema. B cell PU.1 datasets were markedly enriched at genetic risk loci for multiple sclerosis (MS), systemic lupus erythematosus (SLE), and IBD.

Motivated by our prior work implicating Epstein-Barr virus (EBV)-infected B cells in the etiology of diseases such as MS, SLE, and ulcerative colitis^1, 16, 17, 31, 32^, we next evaluated the enrichment of each individual B cell PU.1 dataset across six immune-mediated diseases (**Fig. 4b**). When we stratified B cell ChIP-seq experiments by EBV status, PU.1 datasets from EBV-positive B cells (red) showed substantially stronger and more frequent enrichment at disease risk loci than PU.1 datasets from EBV-negative cells (black). These findings are consistent with a model in which EBV infection reshapes PU.1 binding at disease-associated regions^1, 17^, potentially through cooperation with the EBV transcriptional regulator EBNA2^1, 17, 32^, thereby contributing to gene-environment interactions highlighted by prior molecular and epidemiological studies^33^.

### A compendium of allelic PU.1 binding for benchmarking allelic TF binding prediction methods

To quantify the prevalence of genotype-dependent PU.1 occupancy across hematopoietic contexts, we used the MARIO method^1^ to identify allele-dependent PU.1 binding events in cell lines for which genotype data was available (**Supplementary Data 6**). MARIO identifies allelic TF binding events at heterozygous variants within a ChIP-seq dataset by quantifying the allelic imbalance in read counts overlapping each variant. Across cell types, a large subset of PU.1-bound heterozygous sites displayed significant allelic imbalance (**Fig. 5a**), with the median percentage of allelic PU.1 binding to heterozygotes ranging from 5.3-8.4% across cell types. We next calculated the total number of allele-dependent PU.1 binding events per cell type (**Fig. 5b**). As expected, cell types with larger numbers of datasets and greater overall heterozygosity contributed more allelic events in absolute terms. Nevertheless, all lineages with matched genotype and PU.1 ChIP-seq data yielded hundreds to thousands of allele-dependent sites. These results underscore that genotype-dependent PU.1 occupancy is a widespread feature of the PU.1 DNA binding landscape.

**Figure 5.**
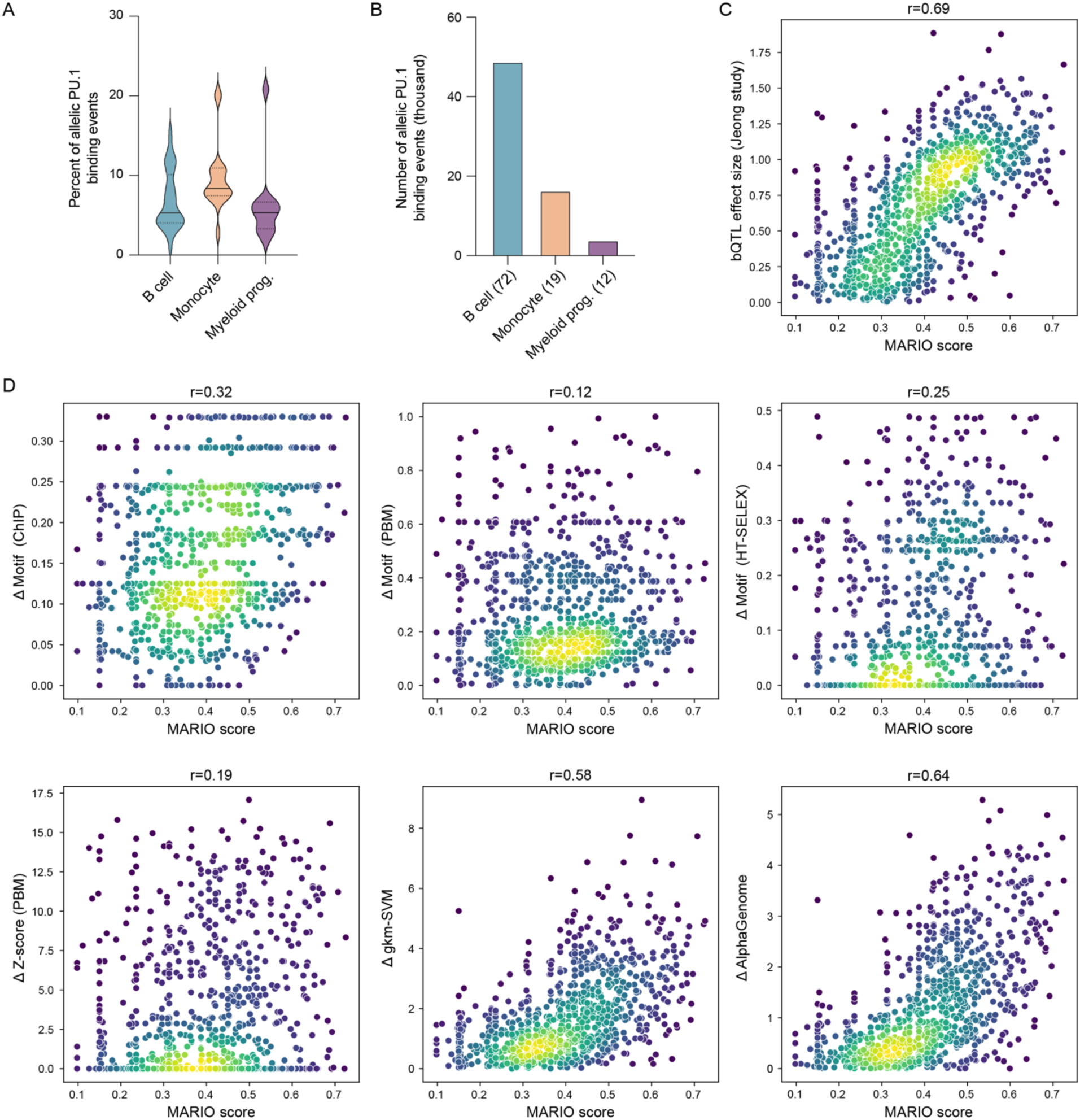
A compendium of PU.1 allelic binding events enables evaluation of methods to predict genotype-dependent TF binding. A. Percent of PU.1-bound heterozygotes with allelic PU.1 binding within each cell type. We identified heterozygotes bound by PU.1 within each ChIP-seq peak set. The percent of significant MARIO results is indicated on the y-axis. The solid line indicates the median and dashed lines indicate quartiles. B. Total number of allele-dependent PU.1 binding events in each cell type. The number of ChIP-seq datasets within each cell type are indicated in parentheses at the bottom. C - D. Evaluation of methods for predicting PU.1 allelic binding using MARIO-derived allelic read imbalance scores at heterozygotes. Each dot represents an evaluation variant. Evaluation variants used are bound by PU.1 in EBV+ B cells and are predicted to alter PU.1 binding sites to varying degrees^15^. Plots indicate density, with yellow indicating higher density and purple lower density. Each panel represents the evaluation of one of seven methods (see Methods). Values at the top of each panel indicate Spearman correlations.

The experimentally-derived allelic imbalance measurements provided by MARIO offer a unique opportunity to benchmark current computational methods for predicting allelic TF binding. Such methods employ different data types and analytical approaches. To this end, we focused on a set of previously identified variants that are bound by PU.1 in EBV+ B cells and are predicted to alter PU.1 binding sites to varying degrees^15^. In this same study, PU.1 binding quantitative trait loci (bQTLs) were also calculated by comparing PU.1 binding genotype-dependent binding strength (ChIP-seq signal) across 49 EBV+ B cell lines derived from individuals with different genetic backgrounds. As expected, the bQTL beta score (a measure of allelic effect size) was strongly correlated with the MARIO allelic reproducibility scores, albeit not perfectly (r = 0.69) (**Fig. 5c, Supplementary Data 7**). These results suggest that MARIO and bQTL analyses capture overlapping but partially distinct sets of genotype-dependent PU.1 binding events, reflecting complementary sensitivities to allelic imbalance versus population-level association signals. Further, they provide an upper-bound performance estimate for evaluating other computational methods based on DNA sequence.

We next used these experimentally derived allelic PU.1 binding events as an empirical benchmark to evaluate sequence-based methods for predicting genotype-dependent TF binding (**Fig. 5d, Supplementary Data 7**). We evaluated three methods based on standard position weight matrix “motifs” (derived from either ChIP-seq, PBM, or HT-SELEX data). We also evaluated three advanced methods based on PBM-derived Z-scores^34^ or machine learning-based approaches based on support vector machines (gkm-SVM^35^) or deep learning-based predictions (AlphaGenome^36^). For each method, we compared the predicted allelic score to the MARIO allelic reproducibility score at each evaluation variant. In general, MARIO-derived allelic imbalance values were correlated with the predictions produced by all methods, indicating that both motif-based and broader sequence-context models capture meaningful, but incomplete, information about allele-dependent PU.1 binding. Notably, of all the methods used to assess allelic PU.1 binding, MARIO and bQTL analysis exhibited the highest correlation (r = 0.69) (**Fig. 5c)**, suggesting that approaches based on experimental ChIP-seq data offer reliable and reproducible identification of genotype-dependent binding events. The limitation of these methods, however, is that significant investment is necessary to obtain these datasets. Computational methods that use large training datasets to build models for predicting TF allelic binding based on DNA sequence alone such as AlphaGenome (r = 0.64) and gkm-SVM (r = 0.58) offer a promising path forward (**Fig. 5d**). Collectively, these analyses define a cross-lineage compendium of allelic PU.1 binding events and illustrate how it can be used to benchmark computational predictors of genotype-dependent TF occupancy.

### Allele-dependent PU.1 binding to disease and cell count associated variants

To illustrate how PU.1 allelic binding links noncoding variation to gene regulation and immune traits, we highlight six loci with robust, reproducible genotype-dependent PU.1 occupancy that are also expression quantitative trait loci (eQTLs) in cells or tissues relevant to immune-mediated disease (**Fig. 6**). At rs3808619, a SLE risk variant, PU.1 strongly preferred the G allele in nineteen PU.1 ChIP–seq datasets in which this SNP was heterozygous. This variant is also a strong bQTL in EBV+ B cells, is predicted by multiple sequence-based computational methods to disrupt PU.1 binding, and is connected to *ZC2HC1A* expression by both activity-by-contact (ABC^37^) scores and blood eQTL data **(Supplementary Data 8 and 9)**. Likewise, we observed consistent allelic PU.1 binding at rs1250568, a PU.1 bQTL associated with MS, together with allelic effects on *PPIF* expression in whole blood. These examples reveal PU.1 allele-dependent binding events to disease-associated variants that can be readily identified by multiple methods.

**Figure 6.**
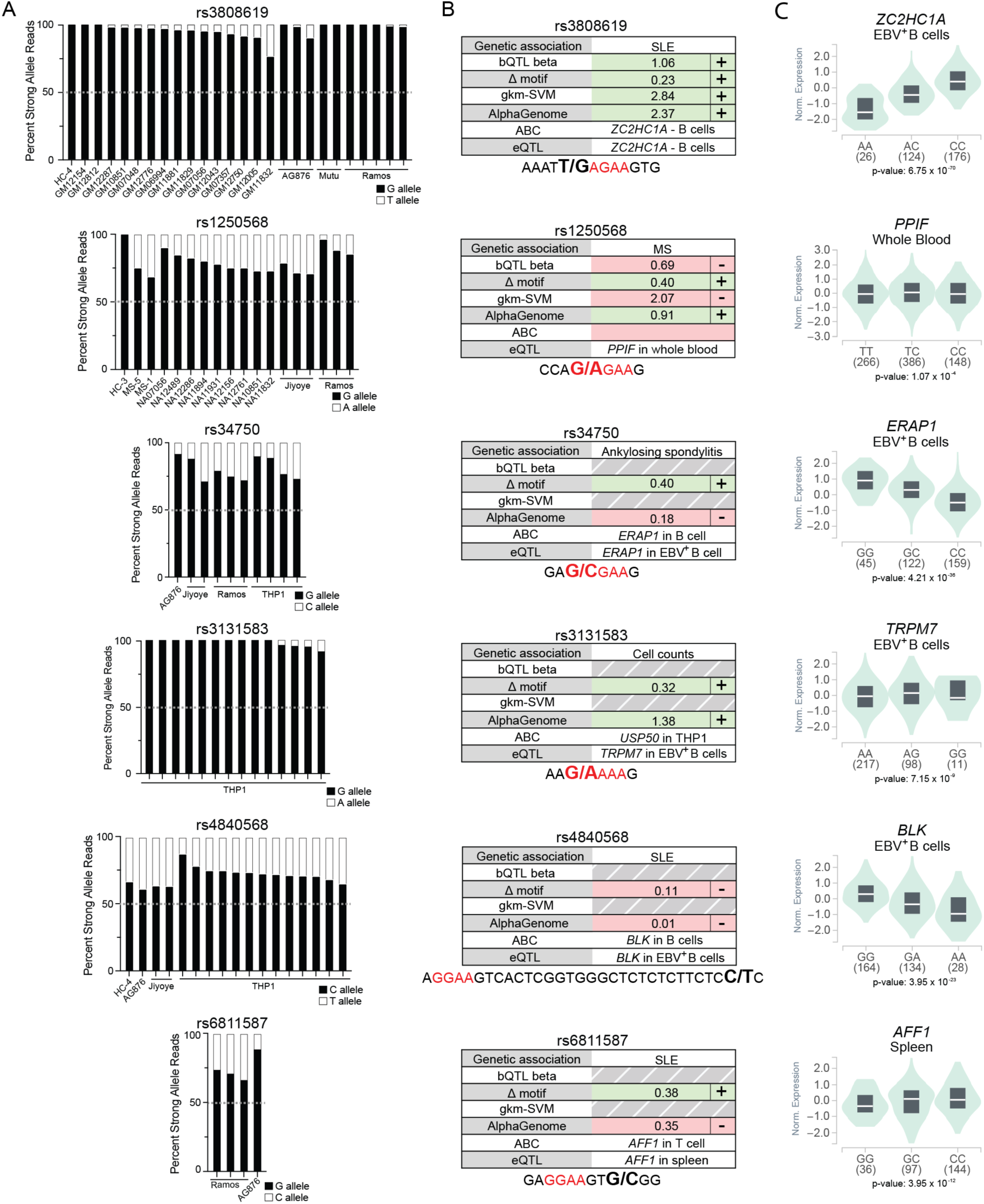
Allele-dependent PU.1 binding to disease and cell count associated variants. Each row illustrates an exemplary genetic variant with robust allelic PU.1 binding and a putative regulatory effect on a nearby gene. A. Percent allelic bias in PU.1 ChIP-seq reads toward the strong base for each allelic PU.1 ChIP–seq dataset. Bars are labeled by cell line; when multiple PU.1 ChIP-seq datasets are available for the same cell line, each dataset is shown separately. B. Additional information for each variant, including the associated immune phenotype, genes linked to the variant by activity-by-contact (ABC) modeling and/or expression quantitative trait loci (eQTLs), and scores from representative allelic computational prediction methods. Positive allelic predictions are colored green (with a “+” sign), negative predictions are colored red (“-“ sign), and missing data are indicated with gray hash marks (see Methods). Scores are oriented so that positive values correspond to the allele predicted to strengthen PU.1 binding (“strong” base). Below each panel, the PU.1-bound DNA sequence surrounding the variant is shown, with the core GGAA consensus motif highlighted in red and the reference and alternate alleles of the SNP indicated at the polymorphic position. C. GTEx eQTL plots displaying the relationship between the genetic variant allele and expression levels of the indicated gene in the indicated tissue.

rs34750 (associated with ankylosing spondylitis and allelic *ERAP1* expression in B cells) and rs3131583 (associated with blood cell counts and allelic *TRPM7* expression in B cells) showed clear genotype-dependent PU.1 binding in MARIO, along with strong sequence-based predictions due to the variant disrupting the core PU.1 motif. However, neither variant was reported as a PU.1 bQTL, likely due to trans effects produced by the different genetic backgrounds of the individuals from which the cells were derived. Similarly, two additional SLE loci, rs4840568 and rs6811587, exhibited robust genotype-dependent PU.1 binding in MARIO. Yet the sequence-based prediction approaches did not prioritize these variants likely because the variants lie adjacent to, rather than within, the core PU.1 binding site. Together, these examples underscore how integrating PU.1 allelic binding data with eQTLs and chromatin-contact maps can reveal allele-dependent disease mechanisms, including putative effector genes. Our results also highlight limitations of current sequence-based models, particularly for variants that act through motif-adjacent or more complex sequence features.

To visualize how allele-dependent PU.1 binding relates to complex trait loci across the genome, we systematically mapped all genomic positions containing allele-dependent PU.1 variants (**Fig. 7, Supplementary Data 10**). The outer track depicts variants with allelic PU.1 binding from our atlas, with the next tracks indicating all variants associated with any disease or blood cell count trait. The inner tracks highlight subsets linked specifically to allergic disease, inflammatory bowel disease, and autoimmune disease, as well as allelic PU.1 sites identified in EBV⁺ and EBV⁻ B cells. Notably, there are many more EBV+ PU.1 allelic binding events to autoimmune risk variants compared to EBV- events, consistent with the established role of EBV in many autoimmune diseases. Across all tracks, allelic PU.1 binding events and trait-associated variants are distributed throughout the genome rather than confined to a limited set of regions. The extensive coverage of all chromosomes and trait categories underscores that the genotype-dependent context of PU.1 occupancy is a pervasive feature of the PU.1 binding landscape, intersecting broadly with loci implicated in immune-mediated and hematologic traits.

**Figure 7.**
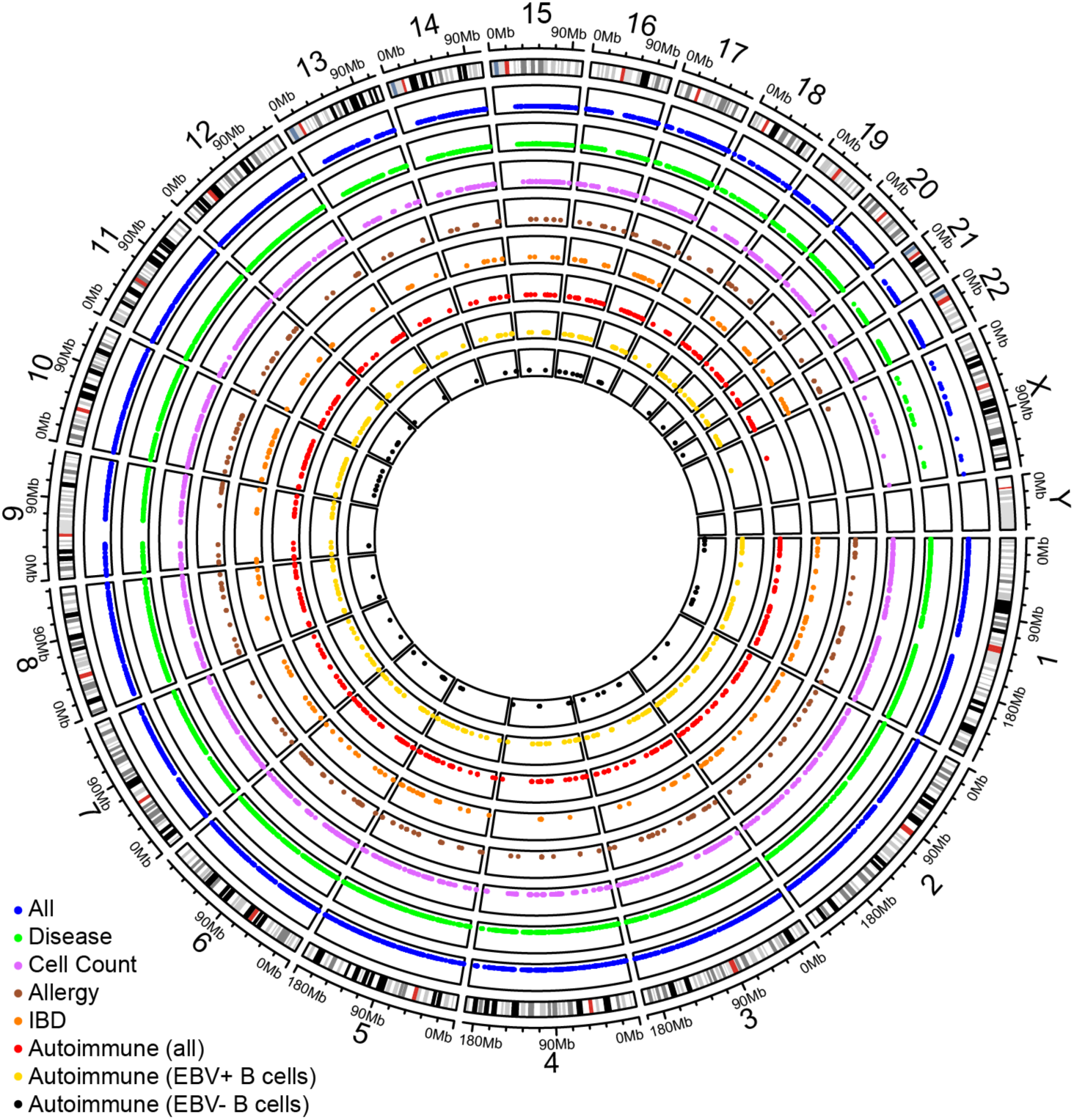
Global map of allelic PU.1 binding at genetic variants. Circos plot depicting human chromosomes (outer circle) with centromeres indicated as a red tick marks. Additional concentric circles represent sets of allelic PU.1 binding events, as follows: blue, all PU.1 allelic binding events; green, PU.1 allelic binding events at disease-associated variants; purple, PU.1 allelic binding events at variants associated with cell count phenotypes; brown, PU.1 allelic binding events at allergy-associated variants; orange, PU.1 allelic binding events at inflammatory bowel disease-associated variants; red, PU.1 allelic binding events at autoimmune disease-associated variants; yellow, PU.1 allelic binding events of autoimmune-disease associated variants identified in EBV-positive B cells; and black, PU.1 allelic binding events of autoimmune-disease associated variants identified in EBV-negative B cells. **Supplementary Data 10** contains a list of loci associated with allelic PU.1 binding.

## DISCUSSION

This work provides a systematic, cross-context view of PU.1 genomic occupancy across human hematopoietic lineages and links this binding landscape to noncoding genetic variation underlying blood cell traits and immune-mediated disease. By assembling and uniformly processing hundreds of PU.1 ChIP-seq datasets spanning primary myeloid and lymphoid cells, tissue-resident macrophages, and EBV-positive/EBV-negative B-cell lines, we show that PU.1 binding profiles recapitulate major hematopoietic identities and cellular states. We identify a large core set of PU.1 binding sites shared across cell types, along with sites that mark cell type-specific regulatory programs. In doing so, the atlas extends single-lineage studies by situating PU.1 within a multi-lineage framework and clarifies how a single lineage-defining TF integrates cellular programs with environmental and genetic contexts to influence immune phenotypes.

Our analyses of DNA sequence features highlight how the short core motif of PU.1 is embedded within a richer grammar of flanking partner transcription factor binding sites that varies across cell types. Shared PU.1 peaks are enriched for broadly used composite motifs, whereas cell-type-restricted peaks show distinctive repertoires consistent with lineage-defining cofactors (e.g., C/EBP in neutrophils, GATA in myeloid progenitor cells, bHLH (TCF4) in B cells, NF-kB in dendritic cells, AP-2 in eosinophils, and FOS/JUN in monocytes). These differences support a model in which PU.1 integrates cell-intrinsic programs with immune signaling through combinatorial binding, including at composite elements^9-11, 26, 38^. They also help explain why the canonical PU.1 motif alone incompletely predicts occupancy and highlights the importance of context-dependent motif combinations in shaping occupancy. Together, our results reinforce a model in which PU.1 functions both on its own and as a collaborator, with binding outcomes dictated by available cofactors, local sequence architecture, and chromatin context.

By intersecting the PU.1 atlas with variants from genome-wide association studies, we identify cellular contexts that are most likely to mediate genetic effects on blood cell traits and immune disease. PU.1 peaks in eosinophils are preferentially enriched at loci associated with eosinophil counts, whereas monocyte and macrophage datasets are enriched at monocyte count loci, and neutrophil datasets are enriched at neutrophil and granulocyte count loci, consistent with a central role for PU.1 in immune cell development^3, 6, 8, 14, 38^. We also observe robust enrichment of PU.1 binding at loci associated with systemic autoimmune diseases, inflammatory bowel disease, and related traits, with distinct patterns across monocyte, macrophage, and B cell datasets.

Environmental context further modulates PU.1 binding at disease-associated regions. Notably, EBV-positive B cell lines show stronger and more extensive enrichment at autoimmune and inflammatory bowel disease loci than EBV-negative cell lines. This is consistent with prior work showing that EBV transcriptional regulators (e.g., EBNA2) co-occupy and rewire host regulatory landscapes at risk loci^1, 32^. This suggests that viral transformation creates a TF environment that accentuates certain risk loci, offering a mechanistic lens for gene-environment interactions implicated by molecular and epidemiological studies^39, 40^.

At the level of genetic context, our atlas reveals that genotype-dependent occupancy is a widespread feature of the PU.1 binding landscape. Assessing ChIP-seq read count allelic imbalance using the MARIO method uncovered thousands of loci with reproducible allelic PU.1 binding events. Many of these sites occur at eQTLs and can be linked to putative effector genes using chromatin contact–based approaches. We present numerous examples to illustrate how allelic PU.1 binding and eQTLs converge to nominate genes as mediators of genetic risk. These include *ZC2HC1A*, *PPIF*, *BLK*, *ERAP1*, *AFF1*, and *TRPM7* as mediators of risk at systemic lupus erythematosus, multiple sclerosis, ankylosing spondylitis, and blood cell count loci. At the same time, our global map of PU.1 allelic binding shows that such variants span the genome and intersect a broad spectrum of immune-related and hematologic traits.

Using these experimentally defined allelic sites, we benchmarked several sequence-based computational methods that aim to predict genotype-dependent TF occupancy. In our analyses, we observe two mechanistic classes of variants: those that directly alter the canonical PU.1 binding site and variants that act through motif-adjacent bases, composite elements, or other types of regulatory syntax. Variants that directly disrupt the canonical PU.1 motif are well captured by both simple motif-based methods as well as advanced methods such as gkm-SVM and AlphaGenome, and often correspond to PU.1 bQTLs. In contrast, many variants with robust allelic binding in our compendium are not prioritized by current prediction frameworks. These observations highlight both the power and the limitations of existing models and argue for next-generation approaches that incorporate broader sequence context, cooperative binding grammar, and cell-type-specific chromatin features when predicting mechanisms impacted by noncoding variation.

Our study establishes a unique resource and conceptual framework for linking context-specific TF binding to human genetic variation across hematopoietic lineages. Further, it reveals how a single TF integrates cellular, environmental, and genetic signals to shape binding and ultimately disease phenotypes. The atlas, allelic binding compendium, and trait-intersection analyses together provide hypotheses about the cellular contexts, target genes, and regulatory mechanisms through which PU.1 contributes to blood cell traits and immune-mediated diseases. More broadly, our TF-centered, cross-context strategy can be applied to other key regulators, especially as large collections of ChIP–seq and chromatin profiling data continue to become available. Creating similar atlases across environmental states (infection, inflammation), developmental trajectories, and cell types will reveal additional principles of cooperative TF binding and disease-specific vulnerabilities. Systematic integration of such resources with genetic, epigenomic, and functional datasets will continue to refine our understanding of how noncoding variation perturbs TF networks to influence human health and disease.

## METHODS

### Cell lines and culture conditions

A list of cell lines used for PU.1 ChIP-seq experiments newly produced for this study is provided in **Supplementary Data 11.** All B cell lines were cultured in Roswell Park Memorial Institute (RPMI) 1640 medium supplemented with 10% fetal bovine serum, 1% antibiotic-antimycotic (Gibco), and 0.2% normocin. The K562 cell line was cultured in RPMI 1640 medium supplemented with 2 mM L-glutamine and 10% fetal bovine serum.

### Isolation of primary human dendritic cells and eosinophils

To isolate primary human dendritic cells, a donor tonsil was first dissociated using a gentleMACS Dissociator (Miltenyi Biotec). A Ficoll-Paque density gradient was used to isolate mononuclear cells from this preparation. A Myeloid Dendritic Cell Isolation Kit (130-094-487, Miltenyi Biotec) was then used to isolate myeloid dendritic cells from this sample, following manufacturer instructions. Human primary eosinophils were isolated from peripheral blood using an Eosinophil Isolation Kit (130-092-010, Miltenyi Biotec) following Ficoll-based granulocyte separation. A total of 7 million eosinophils were used for each ChIP sample.

### PU.1 chromatin immunoprecipitation with sequencing (ChIP-seq)

A total of 34 new ChIP-seq datasets were produced in this study that passed our rigorous quality control standards. Twenty-six were produced in the Kottyan/Weirauch lab (**Supplementary Data 1**). Cells were incubated in a crosslinking solution (1% formaldehyde, 5 mM 4-(2-hydroxyethyl)-1-piperazineëthanesulfonic acid (HEPES) pH 8.0, 10 mM sodium chloride, 0.1 mM ethylenediaminetetraacetic acid (EDTA), and 0.05 mM ethylene glycol tetraacetic acid (EGTA)) in culture medium with 10% fetal bovine serum (FBS) and placed on a tube rotator at room temperature for 10 min. To stop crosslinking, glycine was added to a final concentration of 0.125 M, and the tubes were rotated at room temperature for 5 min. Cells were washed twice with ice-cold phosphate-buffered saline (PBS), resuspended in lysis buffer 1 (50 mM HEPES pH 8.0, 140 mM NaCl, 1 mM EDTA, 10% glycerol, 0.25% Triton X-100, and 0.5% NP-40), and incubated for 10 min on ice. Nuclei were harvested after centrifugation at 5,000 rpm for 10 min, resuspended in lysis buffer 2 (10 mM Tris–HCl pH 8.0, 1 mM EDTA, 200 mM NaCl, and 0.5 mM EGTA), and incubated at room temperature for 10 min. Protease and phosphatase inhibitors (Halt™ Protease and Phosphatase Inhibitor Cocktail (100X), Thermo Fisher Scientific, Waltham, MA) were included in both lysis buffers. Nuclei were resuspended in sonication buffer (10 mM Tris [pH 8.0], 1 mM EDTA, and 0.1% sodium dodecyl sulfate (SDS)). An S220 focused ultrasonicator (COVARIS, Woburn, MA) was used to shear chromatin (150–500-bp fragments) with 10% duty cycle, 175 peak power, and 200 bursts per cycle for 7 min. A portion of the sonicated chromatin was run on an agarose gel to verify fragment sizes. Sheared chromatin was pre-cleared with 10 μL of Protein A Dynabeads (Thermo Fisher Scientific) at 4 °C for 1 h.

Immunoprecipitation of PU.1-chromatin complexes was performed with an SX-8X IP-STAR compact automated system (Diagenode). Beads conjugated to an antibody against PU.1 (Cell Signaling Technology 2266, lot 3) were incubated with precleared chromatin at 4 °C for 8 h. The beads were then washed sequentially with wash buffer 1 (10 mM Tris–HCl [pH 7.5], 150 mM NaCl, 1 mM EDTA, 0.1% SDS, 0.1% NaDOC, and 1% Triton X-100), wash buffer 2 (10 mM Tris–HCl [pH 7.6], 400 mM NaCl, 1 mM EDTA, 0.1% SDS, 0.1% NaDOC, and 1% Triton X-100), wash buffer 3 (10 mM Tris–HCl [pH 8.0], 250 mM LiCl, 1 mM EDTA, 0.5% NaDOC, and 0.5% NP-40), and wash buffer 4 (10 mM Tris–HCl [pH 8.0], 1 mM EDTA, and 0.2% Triton X-100). Finally, the beads were resuspended in 10 mM Tris–HCl (pH 7.5) and used to prepare libraries via ChIPmentation. The ChIP-seq libraries were sequenced as single-end 100-base reads on an Illumina NovaSeq 6000 at the Cincinnati Children’s Hospital Medical Center (CCHMC) Genomics Sequencing Facility, Cincinnati, Ohio.

Eight new ChIP-seq datasets were performed in the Glass lab for this study. In brief, cells were first cross-linked in 2 mM dissuccinimidyl glutarate (Pierce 20593, Thermo Fischer) in PBS for 30 min, followed by subsequent 1% formaldehyde (Sigma) cross-linking in PBS for 10 min at room temperature followed by cross-linked using 1% formaldehyde in PBS for 10 min at room temperature. After cross-linking, glycine (Sigma) was added to a final concentration of 0.2625 M to quench the reaction. Subsequently, cross-linked monocytes were centrifuged (5 min, 1,200 RPM, 4°C), washed twice with PBS, and pellets were snap frozen and stored at −80°C. Frozen cell pellets were resuspended in cell lysis buffer (10 mM HEPES/KOH pH 7.9, 85 mM KCl, 1 mM EDTA, 1.0% IGEPAL CA-630 (Sigma), 1x protease inhibitor cocktail (Roche, Basel, Switzerland), 1 mM PMSF). After 5 min lysis on ice, cells were centrifuged (5 min, 4000 RPM, 4°C), and the supernatant was removed. The pellet was then resuspended in nuclear lysis buffer (10 mM Tris-HCl, pH 8.0, 100 mM NaCl, 1 mM EDTA, 0.5 mM EGTA, 0.1% Na-deoxycholate, 0.5% N-lauroylsarcosine, 1x protease inhibitor cocktail, and 1 mM PMSF) and the chromatin was sheared by sonication on wet ice with a Bioruptor Standard Sonicator (Diagenode, Denville, NJ) for three 15 min cycles each alternating 30 s on and 30 s off on the high setting. Additional Triton X-100 was added to the sonicated chromatin to 10% of the final volume and the lysate was cleared by centrifugation (5 min, 14,000 RPM, 4°C). Input was then saved for subsequent analysis.

Protein A (Invitrogen) pre-bound with antibody was added to the diluted cell lysate overnight at 4°C. Immunoprecipitated complexes were washed three times with 20 mM Tris/HCl pH 7.4150 mM NaCl, 0.1% SDS, 1% Triton X-100, 2 mM EDTA, three times with 10 mM Tris/HCl pH 7.4250 mM LiCl, 1% Triton X-100, 1% sodium deoxycholate, 1 mM EDTA, and two times with Tris-EDTA plus 0.1% Tween-20 before eluting two times with 50 µL elution buffer (TE, 1% SDS, 30 and 10 min, room temperature). Elution buffer was also added to the input. After pooling the eluted samples, the sodium concentration was adjusted to 300 mM and cross-links were reversed overnight at 65°C. Samples were treated with 0.5 mg/ml proteinase K for 1 hr at 55°C and 0.25 mg/ml RNase A for 1 hr at 37°C before DNA was isolated using the ChIP DNA Clean and Concentrator Kit according to the manufacturer’s instructions. For library preparation, NEXTflex DNA barcode adaptors (BioO Scientific, Austin, TX) were ligated to the genomic DNA. Polymerase chain reaction mediated library amplification was performed and final libraries were size selected on 10% TBE gels (Invitrogen).

### PU.1 ChIP-seq data quality control and analysis

296 PU.1 ChIP-seq datasets used in this study were either generated in-house as described above, downloaded from the Gene Expression Omnibus (GEO) on February 20, 2024, or were previously published in PU.1-focused functional genomics studies by Waszak *et al.,* (from LCLs) ^41^, Watt *et al.,* (from neutrophils and monocytes)^14^, Granitto *et al.,* (from LCLs), or Gosselin *et al.,* (from microglia)^42^.

All datasets were uniformly processed. Reads were pre-processed for adapter removal using fastp^43^. Next, the Transcription Factor ChIP-seq Pipeline v2.0 from the ENCODE Project^19-21^ (https://www.encodeproject.org/pipelines/) was used to perform genomic alignments, QC assessments, and peak calling. In brief, ChIP-seq reads were aligned to the human genome (hg38) using Bowtie2 (v. 2.3.4.3)^42^. Aligned reads were then sorted using samtools (v.1.9)^44^ and duplicate reads were removed using Picard^44^ (v. 2.20.7) (https://broadinstitute.github.io/picard/). Peaks were called using the pipeline’s default parameters with MACS2^45^ (v. 2.2.4). ENCODE “exclusion list regions” were removed from the final peak set. All public ChIP-seq datasets (**Supplementary Data 1**) were also analyzed using the same pipeline. The MACS2 peaks generated by the ENCODE pipeline were filtered for q < 0.01.

Next, we uniformly applied a set of rigorous quality control criteria to all 296 ChIP-seq datasets to qualify those included in the study. Datasets were excluded from further analysis for the following reasons: containing fewer than 1,000 high-quality peaks, containing less than 10 million reads after alignment and duplicate read removal, containing fewer than 50% of the expected reads based on published read counts (for public datasets), or containing more than 1% of reads mapping to bacterial genomes in taxonomy analysis with Kraken2^46^. The peaks from the remaining datasets were run through HOMER^27^ to identify the top represented motifs in these peaks, and datasets were removed from further analysis if SPI1/PU.1 was not among the top three motifs. We also performed RELI analysis to identify the pairwise overlap between ChIP-seq peaks; datasets were excluded from further analysis if they had low overlap with any other PU.1 ChIP-seq dataset based on clustering analysis. Following these quality control filters, the 260 remaining high-quality datasets were used in downstream analyses.

### Dimensionality reduction by Uniform Manifold Approximation and Projection (UMAP)

Each PU.1 ChIP-seq peak set was classified into one of ten immune cell types: stem cells, myeloid progenitor cells, neutrophils, monocytes, eosinophils, dendritic cells, macrophages, T cells, B cells, or NK cells (**Supplementary Data 1, Fig. 1A**). The UMAP analysis and all subsequent analyses of PU.1 ChIP-seq peaks excluded T cells and NK cells because none of the datasets in these cell types passed our quality control criteria (they contained very few peaks, which were not enriched for PU.1 motifs), suggesting that PU.1 is not active in these cells.

Using the R package Diffbind^47^ (v. 3.18.0), a set of consensus peaks was generated, and peak summit values were calculated for each PU.1 ChIP-seq peak set. The resulting matrix is equivalent to a high dimensional set of points, where each dimension is represented as a peak and each point represents a peak set, with the peak summit values comprising the data matrix. The UMAP algorithm^48^ was performed on this matrix using the uwot R package (v0.2.3) with default parameters. The first two UMAP components are plotted in Fig. 1B.

### Classification of SPI1 gene expression levels by cell type

We obtained gene expression data from three sources: Human Protein Atlas^22^, Immunological Genome Project^23^, and Database of Immune Cell eQTLs^24^. Because each data source used their own normalization method for reporting gene expression, we qualitatively classified cell types as having high, medium, or low expression of *SPI1*. Qualitative expression levels were widely similar across data sources. The expression category for this study was determined by the first data source with expression based on the following rank: Human Protein Atlas (with protein level normalized expression), Immunological Genome Project, and Database of Immune Cell eQTLs.

### Identification of shared and cell type-specific PU.1 binding sites

We sought to identify PU.1 binding sites that were unique to each cell type, and also those shared between more than one cell type. To obtain a union peak set for each of the ten immune cell types assessed in this study, all peak BED files from PU.1 ChIP-seq datasets passing our quality control criteria were combined within each cell type, and the overlapping peaks were sorted and merged using bedtools^49^ (v. 2.30.0). Intervene^50^ (v. 0.6.5) was run on these union peak sets with the “–save-overlaps” option to generate BED files containing PU.1 peaks specific to each cell type, along with sets of overlapping PU.1 binding sites shared between each combination of two or more cell types. Counts of cell type-specific peaks and shared peaks are plotted in Figure 2A.

### Gene set enrichment analysis using GREAT

The R package rGREAT (v. 2.10.0) was used to perform local GREAT^25^ analysis. GREAT associates ChIP-seq peaks with pathways (e.g., Gene Ontology terms) via a gene proximity-based method^25^. GREAT analysis returns a set of enriched terms with corresponding p-values for each specific PU.1 peak set. These gene ontology terms were intersected with GO slim terms to remove redundancy. The p-values were then readjusted to fit the resulting terms using the Benjamini-Hochberg method.

### Transcription factor DNA binding motif enrichment analysis

Human transcription factor motif enrichment analysis was performed on the cell type-specific PU.1 peak sets using the Hypergeometric Optimization of Motif EnRichment (HOMER) software package^27^. A modified version of HOMER that uses a log base 2 likelihood scoring system was used, and known motifs were obtained from Cis-BP^51^ build 2.0. For PU.1 composite motif analysis, Composite PU.1 motifs were collected using data from the following published studies: Jolma *et al.*, 2015^52^, Xie *et al.*, 2025^29^, Grajales-Reyes *et al.*, 2015^53^, and Heinz *et al*., 2010^27^. These motifs were run using HOMER’s findMotifsGenome function with the "-size given" option against the matched background regions generated with HOMER’s default settings. The threshold for detecting motif presence was set to 70% of the maximum possible score for the given motif. See **Supplementary Datasets 3 and 4** for the full motif enrichment results of these analyses.

### Identification of genetic variants associated with cell count and disease phenotypes

To determine the significance of the overlap between PU.1 ChIP-seq datasets and GWAS-derived disease-associated genetic variants, we generated a custom GWAS catalogue that combines all of the ancestries for a particular disease. To this end, we downloaded the Genome Wide Association Studies Catalogue ^30^ v1.0.3.1, as queried on February 18^th^, 2025. Independent risk loci for each disease/phenotype were identified based on linkage disequilibrium (LD) pruning (*r*2 < 0.2). Risk loci across these independent genetic risk variants were identified by linkage disequilibrium expansion (*r*2 > 0.8) based on 1000 Genomes Data using PLINK (v.1.90b). This created a list of disease risk loci, along with the corresponding genetic variants within the LD block. Finally, the LD expanded list for each ancestry was merged by disease, creating a single list of variants for a given disease. This list of variants was then used for RELI analyses.

### Estimation of the significance of intersected phenotype variants using RELI

The Regulatory Element Locus Intersection (RELI) algorithm^1^ was used to estimate the significance of the overlap between PU.1 binding events and GWAS phenotype variants. As input, RELI takes the genomic coordinates from a set of genomic features. RELI then systematically intersects the coordinates of LD-expanded GWAS Catalog variants with each PU.1 ChIP-seq dataset one at a time, and the number of input regions overlapping the peaks of each dataset are counted. Next, the significance of the intersection of each dataset is calculated (*p*-value) using a simulation-based procedure in which the peaks from the input dataset are randomly distributed within the union coordinates of open chromatin from human cells. A distribution of expected overlap values is then created from 1,000 iterations of random sampling from this negative set, each time choosing a set of negative examples that match the input set in terms of the total number of genomic loci. The distribution of the expected overlap values from the randomized data resembles a normal distribution and is thus used to generate a Z-score and corresponding *p*-value estimating the significance of the observed number of input regions that overlap each dataset. *P*-values are corrected using Bonferroni’s method. This procedure was completed for all datasets in the PU.1 ChIP-seq library.

Once *p*-values were calculated for each dataset-phenotype pair, they were aggregated by immune cell classification (e.g. B cell) using Fisher’s method. GWAS phenotypes were manually curated into two categories: cell count and disease. For each category, a matrix was created where immune cell categories were the rows, phenotypes were the columns, and aggregated *p-*values were the values.

### Whole genome sequencing and variant calling

DNA for experiments performed in our laboratory was isolated using PureLink Genomic DNA Kit (ThermoFisher). Whole genome sequencing was performed using DNBseq next generation sequencing technology. Libraries were sequenced on an Illumina NovaSeq to generate 100-base, paired-end reads. Sequencing reads were aligned, and variant calls variants were called with the Genome Analysis Toolkit (GATK) Unified Genotyper following the GATK Best Practices 3.3^54-56^. Genotypes obtained from sequences from the 1000 genomes project were used for experiments performed in Waszak *et al*.,^41^. To identify heterozygous variants, we identified variants within each individual dataset that had an allele frequency greater than 40% using PLINK^55^ v2.0. These heterozygous loci were used in MARIO analyses (see below).

### Identification of allele-dependent sequencing reads using MARIO

Allele-dependent behavior was identified in sequencing reads using the MARIO pipeline^1^. Briefly, MARIO identifies allele-dependent behavior by weighing the imbalance between the number of reads that are mapped to each allele and the total number of reads mapped at each heterozygous genetic variant. These variables are combined into a single Allelic Reproducibility Score (ARS), which reflects the degree of allelic behavior observed for the given heterozygous variant in the given data set. For heterozygous variants overlapped by at least 7 PU.1 ChIP-seq reads, MARIO ARS values (“MARIO score”) exceeding 0.4 were considered to be allelic, following our previous study^1^.

### Evaluation of methods for predicting allelic PU.1 binding

We used allele-dependent PU.1 binding events identified by MARIO to evaluate a panel of computational methods for predicting allelic TF binding. To this end, we focused on a set of previously identified variants that are bound by PU.1 in EBV+ B cells and are predicted to alter PU.1 binding sites to varying degrees^15^. We restricted this set to variants that are heterozygous and located inside a peak in at least one of the EBV+ B cell lines with available PU.1 ChIP-seq data. For variants meeting these criteria in multiple datasets, we used the median MARIO score in the evaluations. This final set of 1,099 MARIO allelic scores was used to evaluate a panel of methods for predicting allelic TF binding. For each of these evaluation variants, we predicted allelic PU.1 binding using seven different methods: (1) bQTLs in EBV+ B cells obtained from Jeong *et al*.,^15^ (2,3,4) position frequency matrix “motifs” derived from PBM, HT-SELEX, or ChIP-seq experiments obtained from the Cis-BP database^51^ (Cis-BP accessions M00210_3.00, M03169_3.00, and M08402_3.00); (5) PBM Z-scores for experiment “Sfpi1_1034” from Badis *et al*.,^57^; (6) gkm-SVM, a machine learning method employing gapped k-mers^35^; and (7) the “ChIP-TF variant scorer” component of AlphaGenome, a recently developed deep learning method^36^. For each method, a “delta score” was calculated quantifying the predicted difference in binding between the two alleles. For motif-based predictions, each variant allele was scored with the given motif using the standard log-likelihood scoring system^58^, and the resulting scores were converted to “percent maximum” scores by dividing the value by the highest possible value obtained by the motif and mapping negative values to zero. For Z-score-based predictions, the allele sequences were broken down into their constitutive overlapping 8mers, the highest 8mer Z-score was recorded for each allele, and the difference between the best Z-score of each allele was calculated, following previous methods^59^. The allelic PU.1 binding predictions produced by each method were then evaluated by comparing to MARIO allelic scores using the Spearman correlation. In **Figure 6**, “positive” allelic events were defined as those with scores at least one standard deviation above the mean, where the mean and standard deviation were determined from the distribution of scores from all predictions for the given method.

## Data Availability

All data produced in the present study are available upon reasonable request to the authors

## SUPPLEMENTAL FIGURES

**Supplemental Figure 1. Relationship of PU.1 ChIP-seq peaks by antibody, experiment of origin, or peak count.** The UMAP from **Fig. 1b** is reproduced here, with each point representing a PU.1 ChIP-seq experiment, and the points colored by A) antibody, B) experiment of origin, or C) peak count range. Experiments are clustered using UMAP based on read counts within each peak.

## SUPPLEMENTARY DATA

**Supplementary Data 1.** PU.1 ChIP-seq metadata and quality control metrics

**Supplementary Data 2.** Enrichment of Gene Ontology (GO) terms in genes associated with PU.1 ChIP-seq peaks using GREAT

**Supplementary Data 3.** Enrichment of transcription factor binding motifs at PU.1 ChIP-seq peaks relative to background sequences

**Supplementary Data 4.** Enrichment of PU.1 composite motifs at PU.1 ChIP-seq peaks relative to background sequences

**Supplementary Data 5.** RELI analysis computing the overlap between PU.1 ChIP-seq datasets and phenotype-associated variants from the GWAS Catalog

**Supplementary Data 6.** Genotype-dependent PU.1 binding events identified using MARIO

**Supplementary Data 7.** Evaluation scores for PU.1 binding

**Supplementary Data 8.** Significant variant-gene associations in GTEx v10, for all variants with allelic PU.1 binding and at least one phenotype association

**Supplementary Data 9.** Activity-By-Contact (ABC) variant-gene annotations (Nasser *et al*., *Nature* 2021) in immune cells for variants from Figure 6.

**Supplementary Data 10.** hg38 coordinates of variants allelically bound by PU.1 in multiple conditions, used to create Circos plots

**Supplementary Data 11.** Cell lines used for PU.1 ChIP-seq in this study

## ACKNOWLEDGMENTS

Schematic figures were made in BioRender. We acknowledge the support of the Informatics Shared Facility in Information Services for Research (IS4R) at Cincinnati Children’s Hospital Medical Center (RRID: SCR_022622). We also acknowledge Cincinnati Children’s Hospital Medical Center Genomics Sequencing Facility (RRID: SCR_022630) for high-throughput sequencing. We thank Kevin Ernst for assistance with database management and computational support. We thank members of the Division of Pediatric Otolaryngology for their longstanding collaboration to obtain tonsils for research under an IRB-approved protocol with informed consent, including Angie Duggins and Douglas von Allmen. We thank the following members of the Waggoner lab for experimental support: Owen Clay, Stacey Cranert, Durga Krishnamurthy, David Ohayon, and Fazeela Yaqoob.

## AUTHOR CONTRIBUTIONS

L.C.K., M.T.W., and S.N.W. conceived of the study. L.C.K., M.T.W., D.A.L., M.S.S., and L.P.L. directed the data analysis. D.A.L., M.S.S., L.P.L., S.P., X.C., L.E.E., D.A.R.-T., A.V.H., and P.J.D performed the data analysis. L.C.K., M.T.W., S.N.W., C.K.G., T.D.T., and M.E.R. directed the experiments. C.Y., C.R.F., J.M.F., A.A.D., K.A.D., J.S.S., and D.F.S. obtained samples and/or performed experiments. L.C.K., M.T.W., D.A.L., M.S.S., and L.P.L. prepared the manuscript and figures. L.E.E. publicly deposited the data.

