## Supplementary figures and images for "Comparison of PU.1 genomic binding across immune cells reveals cell type-specific roles in autoimmune disease"

### Supplemental Figure 1

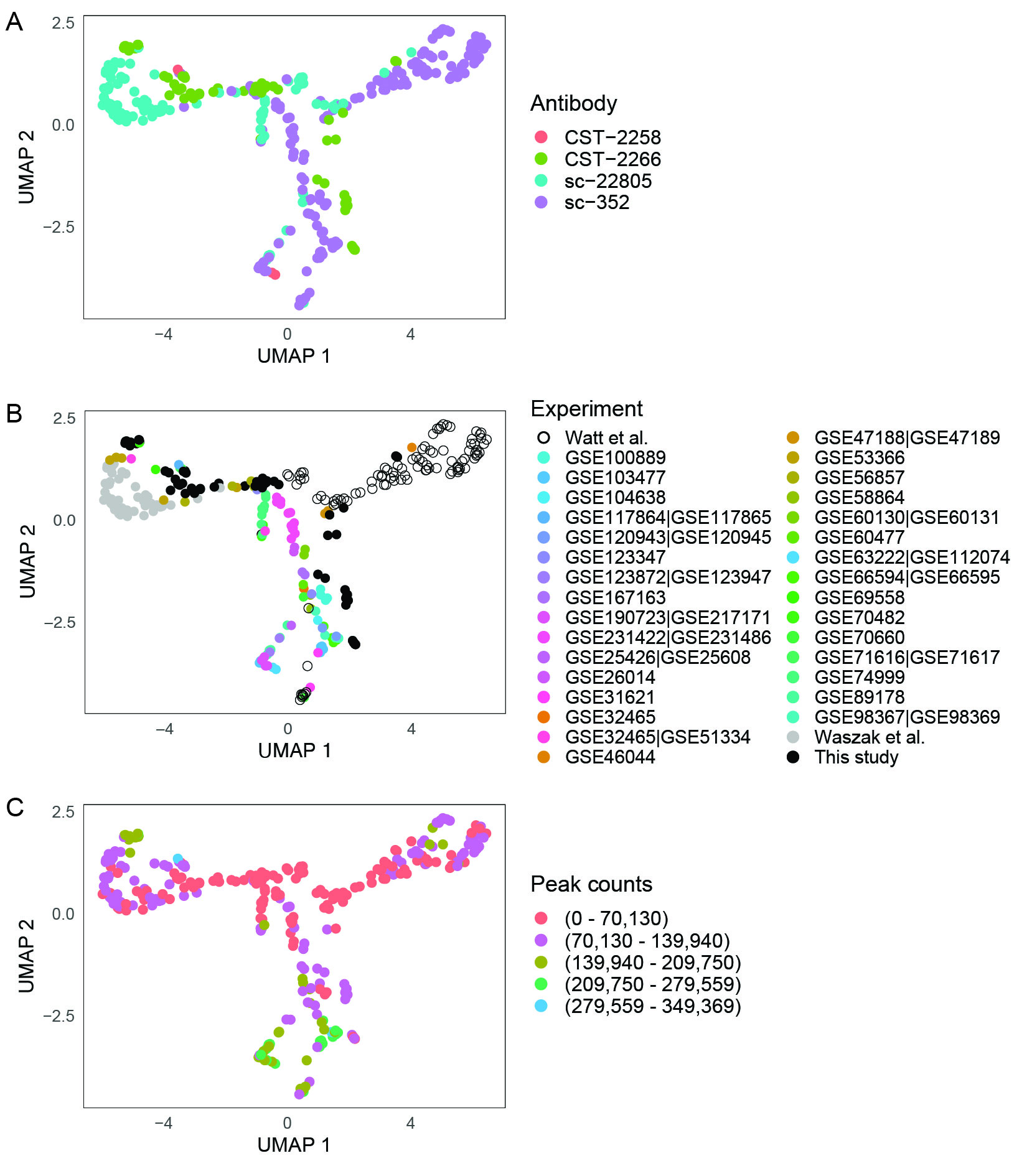
